# From aging to Alzheimer’s disease: concordant brain DNA methylation changes in late life

**DOI:** 10.1101/2025.06.17.25329345

**Authors:** David Lukacsovich, Juan I. Young, Lissette Gomez, Michael A. Schmidt, Wei Zhang, Brian W. Kunkle, X. Steven Chen, Eden R. Martin, Lily Wang

## Abstract

Aging is the strongest risk factor for Alzheimer’s disease (AD), yet the molecular mechanisms linking aging to AD remain poorly understood. DNA methylation (DNAm) is an epigenetic modification that plays a critical role in gene regulation and has been implicated in both aging and AD. In this study, we performed a meta-analysis of DNAm profiles in the prefrontal cortex using two large, independent postmortem brain cohorts, the Religious Orders Study and Memory and Aging Project (ROSMAP) and Brains for Dementia Research (BDR), to identify DNAm differences associated with aging in late life.

We identified 3,264 CpGs significantly associated with aging, the majority of which were hypermethylated and enriched in promoter regions and CpG islands. These aging-associated DNAm changes were significantly overrepresented in genes involved in immune regulation and metabolic pathways. When compared with AD-associated DNAm changes, we found a significant overlap, with nearly all CpGs and differentially methylated regions (DMRs) that were associated with both aging and AD Braak stage displaying concordant directionality. This supports the hypothesis that aging and AD are interconnected at the molecular level.

Further integrative analyses indicated that a number of these DNAm variants may have functional relevance in AD. By integrating blood DNAm data, we identified multiple CpGs that showed significant brain-to-blood correlations and were involved in both aging and AD pathogenesis. Co-localization analyses with genome-wide association study (GWAS) data revealed shared genetic regulation of DNAm and dementia at several AD risk loci. Out-of-sample validation using the Alzheimer’s Disease Neuroimaging Initiative (ADNI) dataset demonstrated that, among 334 CpGs showing concordant DNAm changes in aging and AD, baseline DNAm levels at cg10752406 in the *AZU1* promoter were significantly associated with AD progression at a 5% false discovery rate, even after adjusting for age, sex, *APOE* ε4 allele status, years of education, and baseline MMSE. Notably, this CpG also showed a strong brain–blood DNAm correlation, further supporting its potential as a peripheral biomarker for AD. Our study provides valuable insights into the epigenetic landscape of aging and its implications for AD, suggesting that aging-related epigenetic modifications may provide a viable source of biomarkers for AD.

## INTRODUCTION

Alzheimer’s disease (AD), the most common neurodegenerative disorder, is anticipated to affect nearly 13 million people by 2050^1^. This rise in AD cases is expected to increase healthcare costs significantly, from $355 billion in 2021 to $1.1 trillion in 2050. Aging is the strongest risk factor for AD. The percentage of people with AD increases from 5% for 65–74-year-olds to 14% in 75-84-year-olds and 35% in people older than 85 ^2,3^. However, our current understanding of how age-related molecular processes contribute to AD is still very limited.

Epigenetic modifications, particularly DNA methylation (DNAm), play a crucial role in understanding aging and AD. It has been observed that as people age, DNAm decreases at intergenic regions but increases at many promoter-associated CpG islands regions^4^. Common age-associated changes across many individuals have been observed at many loci^5-13^. DNAm also plays an important role in AD^14-18^. In Zhang et al. (2020), we identified methylation differences that were consistently associated with AD neuropathology in multiple brain EWAS^19^. Our analyses showed the majority of these AD neuropathology-associated DNAm differences in the brain were hyper-methylated with advancing AD Braak stage, many are located in CpG islands, and were significantly enriched in polycomb repressive regions and genes involved in the immune processes ^19^.

Although DNAm in aging has been previously described, their relevance to AD is less explored. Clearly, aging and AD are intertwined ^20^, and the exact contribution of age-related DNAm changes to AD remains to be clarified. Most previous studies of DNAm differences in AD treated age as a confounding variable to be adjusted for and did not discuss how AD-associated DNAm differences relate to those occurring in normal aging. In addition, while a recent study identified distinct risk factors for dementia in early (< 45 years), midlife (45-60 years), and late-life (≥ 65 years) ^21^, most DNAm studies in aging did not stratify by age group of the subjects in a particular period of life. Finally, most of the aging studies have utilized blood samples. However, for neurological disorders such as AD, the use of disease-relevant tissue (e.g., brain tissues) is often preferred for epigenetic studies.

In this study, we aimed to compare the changes in brain DNAm during normal aging with those occurring in AD in late life, a time when AD is most prevalent. To this end, we performed a meta-analysis of two large cohorts of postmortem prefrontal cortex brain samples from subjects over 65 years old, generated by the Religious Orders Study and Rush Memory and Aging Project (together as ROSMAP^22^) and the Brains for Dementia Research (BDR) study^23^. By comparing the age-associated DNAm changes with those previously identified in AD, we identified a subset of DNAm alterations that are common in normal aging and AD. Next, to evaluate the feasibility of these DNAm changes as biomarkers, we performed colocalization analysis incorporating results from recent AD GWAS, assessed brain-to-blood DNAm correlations at these CpGs, and performed an out-of-sample validation using an external longitudinal AD progression dataset. Our findings offer valuable insights into the DNAm changes in aging and their implications for AD.

## METHODS

### Study cohorts

Our meta-analysis included a total of 332 DNA methylation profiles from prefrontal cortex brain samples in two independent cohorts, the ROSMAP^22^ and BDR ^24^ studies. These datasets can be accessed at the AD Knowledge Portal (accession: syn3157275) and GEO database (accession: GSE197305). In the ROSMAP dataset, we excluded brain samples from black subjects (n = 2) or subjects with Hispanic origin (n = 2). We additionally excluded 4 samples with postmortem interval greater than 72 hours ^25^.

### Pre-processing of DNA methylation data

The samples in the ROSMAP and BDR study were measured using the Infinium HumanMethylation450 and Infinium MethylationEPIC BeadChip, respectively. Supplementary Table 1 shows the number of CpGs and samples removed at each quality control step. Quality control for CpG probes included removing probes with detection *P-*value > 0.01 in any sample in a cohort using the minfi R package, removing probes that are cross-reactive ^26^, having a single nucleotide polymorphism (SNP) with minor allele frequency (MAF) ≥ 0.01 present in the last 5 base pairs of the probe, or located on the sex chromosomes using the DMRcate R package (with function rmSNPandCH and parameters dist = 5, mafcut = 0.01).

For quality control of samples, we removed samples with low bisulfite conversion efficiency (i.e., < 85%) or those with discrepancies between the recorded sex and DNAm-predicted sex status. The quality-controlled methylation datasets were then subjected to the QN.BMIQ normalization procedure ^27^. Specifically, we first applied quantile normalization as implemented in the lumi R package to remove systematic effects between samples. Next, we applied the β-mixture quantile normalization (BMIQ) procedure ^28^ as implemented in the wateRmelon R package^29^ to normalize beta values of type 1 and type 2 design probes within the Illumina arrays. Finally, we performed principal component analysis (PCA) analysis on the normalized data using the 50,000 most variable CpGs for each cohort. Samples that deviated more than ± 3 standard deviations from the mean of PC1 and PC2 were removed.

In the BDR dataset, the ages of the brain samples were estimated using the cortical clock method (https://github.com/gemmashireby/CorticalClock), which was shown to accurately predict age in the human brain cortex for BDR samples ^23^. To select brain samples with minimal tau pathology, we used a clustering algorithm. Previously, Shireby et al. (2022) developed DNA methylation reference panels specific to various cell types, including NeuN+ (neuronal-enriched), SOX10+ (oligodendrocyte-enriched), and NeuN–/SOX10– (enriched in microglia and astrocytes) ^24^. These panels, along with the Houseman deconvolution algorithm ^30^, were used to estimate relative cell-type proportions in bulk cortex samples from the BDR cohort. It was observed that an increase in tau pathology significantly correlated with a decrease in the proportion of NeuN+ (neuronal) cells, a reduction in NeuN-/SOX10- (microglial/astrocytic) proportions, and an increase in SOX10+ (oligodendrocytic) proportions in the brain frontal cortex ^24^. To select brain samples with Braak stages 0-2, we applied the k-means clustering algorithm (with k = 3) to the estimated cell-type proportions for the BDR brain samples from Shireby et al. (2022) ^24^ (Supplementary Figure 1). We identified 196 BDR brain samples with Braak stages 0-2, which matches the sample size shown in Table 1 of Smith et al. (2021) ^31^.

**Table 1.** Top 10 most significant CpGs associated with age at death in meta-analysis of ROSMAP and BDR datasets. For each CpG, annotations include the location of the CpG (Chr, Position), nearby genes based on GREAT software. The inverse-variance weighted meta-analysis regression model results include estimated effect size (estimate) where CpGs that are hyper-methylated with increased age have positive values, standard error (stdErr), P-value (pValue), and false discovery rate (fdr) for multiple comparison corrections. The last column (direction) indicates the direction of effects in ROSMAP and BDR cohorts. All P-values are two-sided. In GREAT annotation, the numbers in parentheses indicate distance from the TSS.

| CpG | Annotations |  |  | Meta-alysis results |  |  |  |  |
| --- | --- | --- | --- | --- | --- | --- | --- | --- |
|  | chr | position | GREAT_annotation | estimate | stdErr | pValue | fdr | direction |
| cg21572722 | chr6 | 11044894 | ELOVL2 (-347) | 0.018 | 0.002 | 7.04E-31 | 5.21E-25 | ++ |
| cg24724428 | chr6 | 11044888 | ELOVL2 (-341) | 0.031 | 0.003 | 1.11E-20 | 4.11E-15 | ++ |
| cg20692569 | chr7 | 72848481 | FZD9 (+372) | 0.028 | 0.003 | 2.33E-20 | 5.66E-15 | ++ |
| cg13814485 | chr10 | 8095500 | GATA3 (-1156) | 0.037 | 0.004 | 3.06E-20 | 5.66E-15 | ++ |
| cg07544187 | chr19 | 19651235 | CILP2 (+2161);PBX4 (+78490) | 0.041 | 0.005 | 1.36E-19 | 2.01E-14 | ++ |
| cg09232937 | chr5 | 3595970 | IRX1 (-198) | 0.026 | 0.003 | 2.58E-18 | 3.18E-13 | ++ |
| cg18279094 | chr1 | 63790044 | ALG6 (-43217);FOXD3 (+1314) | 0.040 | 0.005 | 4.06E-17 | 4.29E-12 | ++ |
| cg00495775 | chr2 | 176972041 | HOXD10 (-9266);HOXD11 (+27) | 0.020 | 0.002 | 6.14E-17 | 5.68E-12 | ++ |
| cg22331349 | chr19 | 52391350 | ZNF577 (-146) | 0.020 | 0.002 | 2.21E-16 | 1.81E-11 | ++ |
| cg16867657 | chr6 | 11044877 | ELOVL2 (-330) | 0.017 | 0.002 | 2.51E-16 | 1.86E-11 | ++ |

### Statistical analysis of individual CpGs

First, we performed cohort-specific analyses for individual CpGs. The association between CpG methylation levels and age at death was assessed using linear statistical models in each cohort. Given that methylation M-values (logit transformation of methylation beta values) have better statistical properties (i.e., homoscedasticity) for linear regression models, we used the M-values as the outcome variable in our statistical models. For both the ROSMAP and BDR cohorts, we adjusted for potential confounding factors including sex, estimated cell-type proportions (i.e., proportions of neurons) in the samples, as well as batch effects. For the BDR dataset, we did not have access to postmortem interval (PMI) information. In the ROSMAP dataset, PMI was not significantly associated with age at death (correlation = 0.0033, *P-*value = 0.9691), thus unlikely to be a confounder. We therefore did not include PMI as a covariate in the model.

### Inflation assessment and correction

We estimated genomic inflation factors (lambda values) using the bacon method ^32^, which was specifically proposed for accurate assessment and correction of inflation in epigenome-wide association studies (EWAS). In the analysis of individual CpGs, the lambda values (λ) were 1.12 and 1.18 for the ROSMAP and BDR cohorts, respectively. The estimated bias for these cohorts were -0.13 and -0.17, respectively. After inflation correction using the bacon method, the estimated bias was 1.05 × 10^-3^ and -3.40×10^-4^, and the estimated inflation factors were λ = 1.00 and 1.00 for the ROSMAP and BDR cohorts, respectively. For the analysis of regions, before correction, the inflation estimates were λ = 1.14 and 1.23, and the bias estimates were -0.19 and -0.09 for the ROSMAP and BDR cohorts, respectively. After bacon correction, the inflation estimates were 1.00 and 1.01, and the estimated biases were -2.33 ×10^-3^ and 1.34 ×10^-2^ for these two cohorts.

### Meta-analysis

The *bacon* method was also used to obtain inflation-corrected effect sizes, standard errors, and *P-*values for each cohort, which were then combined by inverse-variance weighted meta-analysis models using R package meta. To account for multiple comparisons, False Discovery Rate (FDR) based on the method of Benjamin and Hochberg ^33^ were computed to adjust bacon-corrected *P-*values. We considered individual CpGs with 5% FDR and with effect sizes in concordant directions in both cohorts to be statistically significant.

### Region-based analysis

To identify differentially methylated regions significantly associated with age at death, we used two analytical approaches, the comb-p ^34^ approach and the coMethDMR ^35^ approach, and selected significant DMRs identified by both methods. Briefly, comb-p takes single CpG *P-*values and locations of CpG sites to scan the genome for regions enriched with a series of adjacent low *P-*values. In our analysis, we used meta-analysis *P*-values of the two brain sample cohorts as input for comb-p. We used the default parameter setting for our comb-p analysis, with parameters --seed 1e-3 and --dist 200, which required a *P-*value of 10^-3^ to start a region and extend the region if another *P-*value was within 200 base pairs. As comb-p uses the Sidak method ^36^ to adjust for multiple comparisons, we selected DMRs with Sidak *P-*values less than 0.05. In addition, we further required the selected DMRs to have at least 3 CpGs, and a consistent direction of change across all CpGs mapped within the region.

In the coMethDMR approach, the “contiguous genomic regions” are genomic regions on the Illumina array covered with clusters of contiguous CpGs where the maximum separation between any two consecutive probes is 200 base pairs. First, coMethDMR selects co-methylated sub-regions within the contiguous genomic regions. Next, we summarized methylation M values within these co-methylated sub-regions using medians and tested them against age at death, adjusting for sex, estimated cell-type proportions, and batch effects using linear regression models. The bacon method ^32^ was next applied to cohort-specific coMethDMR test statistics to obtain inflation-corrected effect sizes, standard errors, and *P-* values, which were then combined by inverse-variance weighted meta-analysis models using R package meta. We considered co-methylated DMRs with at least 3 CpGs and meta-analysis FDR < 0.05 to be significant in the coMethDMR analysis. Finally, we selected significant DMRs identified by both comb-p and coMethDMR approaches for subsequent analyses.

### Functional annotations and enrichment analyses

The significant DNAm differences at individual CpGs and DMRs were annotated using the Illumina annotations and the GREAT (Genomic Regions Enrichment of Annotations Tool) software^37^ with default parameters. We tested for significant over- and under-representation of various genomic features (island, open sea, shelf, shore, 1stExon, 3’UTR, 5’UTR, body, intergenic, TSS1500, TSS200) and chromatin states among the DNAm differences using two-sided Fisher’s exact test. The chromatin states (core 15-state model estimates) were obtained from the Roadmap Epigenomics Project^38^ website (https://egg2.wustl.edu/roadmap/web_portal/chr_state_learning.html#core_15state), which were previously estimated with ChromHMM^39^ using a DLPFC tissue sample (E073). Given a total of 26 genomic features (11 genomic locations and 15 chromatin states), we considered *P-*values less than 1.92 × 10^-3^ (i.e., 0.05/26) to be statistically significant.

For each genomic feature, Fisher’s exact test was used to compare the proportions of CpGs mapped to a particular type of genomic feature (e.g., CpG island) among significant CpGs (foreground) vs. the proportion of CpGs mapped to the same genomic feature in all the tested CpGs (background). Similarly, for region-based analysis, Fisher’s exact test compared the proportion of CpGs within significant DMRs that mapped to a particular type of genomic region (e.g., CpG islands) (foreground) to the proportion of CpGs in contiguous genomic regions on the array (described under “Region-based analysis” above) that mapped to the same type of genomic region (background).

In addition, the LOLA ((Locus Overlap Analysis) software^40^ was used to identify transcription factors and chromatin proteins with binding sites significantly enriched with aging-associated CpGs and DMRs. Finally, to identify biological pathways enriched with significant methylation changes, we combined significant individual CpGs and CpGs from DMRs, given that significant DMRs and individual CpGs can co-localize to the same gene, and tested for enrichment using the missMethyl R package^41^.

### Comparison with AD Braak stage-associated DNA methylation differences

The significant AD Braak stage-associated CpGs and DMRs were obtained from Supplementary Data 15 and 2, respectively, in Zhang et al. (2020) ^19^. The significant results from the matched samples analysis were obtained from Supplementary Data 6 and 7 in Zhang et al. (2020) ^19^. The enrichment analysis results of AD Braak stage-associated CpGs and DMRs were from Supplementary Data 3 and 4 in Zhang et al. (2020). Both the analyses by Zhang et al. (2020) and this manuscript used the same DNAm data processing and inflation correction procedures, making the results highly comparable.

### Correlation and co-localization with genetic susceptibility loci

We searched brain mQTLs of significant aging-associated DNAm at individual CpGs and CpGs located within significant DMRs using the xQTL database ^42^, which includes Bonferroni-corrected associations (http://mqtldb.godmc.org.uk/downloads). CpG to SNP associations were considered *cis* if the associations between SNPs and DNA methylation CpGs were within a 5Kb window for mQTL analysis. Genome-wide summary statistics for genetic variants associated with dementia, as described in Bellenguez et al. (2022)) ^43^, were obtained from the European Bioinformatics Institute GWAS Catalog (https://www.ebi.ac.uk/gwas/) under accession no. GCST90027158. Colocalization analysis was performed using the coloc R package with default parameters.

### Correlations between DNA methylation in blood and brain samples

Using the London cohort, which consisted of 69 samples with match PFC and blood samples^19^, we computed Spearman correlations of blood and brain DNAm levels at significant individual CpGs and CpGs located in significant DMRs using an adjusted analysis. We accounted for confounding factors, including estimated neuron proportions for brain samples (or estimated blood cell-type proportions for blood samples), batch effects, age at death (for brain samples) or age at blood draw (for blood samples), and sex. Specifically, these confounding effects were removed by first fitting linear regression models (methylation *M* value ∼ age + sex + batch + cell-type proportions) separately for brain and blood samples, and extracting the residuals. Spearman correlations were then computed using these residuals.

### Validation of concordant brain DNAm differences in aging and AD

To compare our results with previous reported findings, we used the CpG Query tool in the MIAMI-AD database^44^ (https://miami-ad.org/) to examine the 334 significant CpGs concordantly associated with both aging and AD. For the phenotype input, we selected “AD Biomarker”, “AD Neuropathology”, “Dementia Clinical Diagnosis”, and “Mild Cognitive Impairment”.

In addition, we evaluated the predictive value of these concordant CpGs for disease progression, by performing an out-of-sample validation using the ADNI dataset. These analyses were performed using the coxph function in the survival R package. Disease progression was defined as the conversion from CU to MCI or AD, and from MCI to AD, and the models adjusted for multiple covariates: Surv (Conversion event, follow-up time) ∼ cpg.beta.value + age + sex + APOE ε4 status + years of education + baseline MMSE score. The Kaplan-Meier curves were generated using the survminer R package.

## RESULTS

### Study cohort characteristics

The average ages at death for the ROSMAP and BDR cohorts were 83 ± 6.09 and 80 ± 7.44 years, respectively (Supplementary Table 2). About 51% of the subjects in each cohort were males. To study DNA methylation associated with normal aging in late life, our study focused on brain samples from subjects older than 65 years. Moreover, we carefully selected brain samples with neurofibrillary tangle (NFT) Braak stages ranging from 0 to 2, indicating minimal or absent AD neuropathology.

### DNA methylation differences are significantly associated with aging at individual CpGs and co-methylated regions

Adjusting for estimated cell-type proportions (i.e., the proportion of neurons), sex, and batch effects, our meta-analysis of individual CpGs identified 3264 significant age-associated CpGs at a 5% false discovery rate (FDR) and with a consistent direction of change in both ROSMAP and BDR cohorts (Figure 1, Supplementary Table 3). Among them, 394 CpGs also reached the 9 × 10^-8^ genome-wide significance level^45^. Consistent with previous studies ^46,47^, we observed the majority of the significant CpGs (84.83%, 2769 CpGs) were hypermethylated with an increase in age.

**Figure 1.**
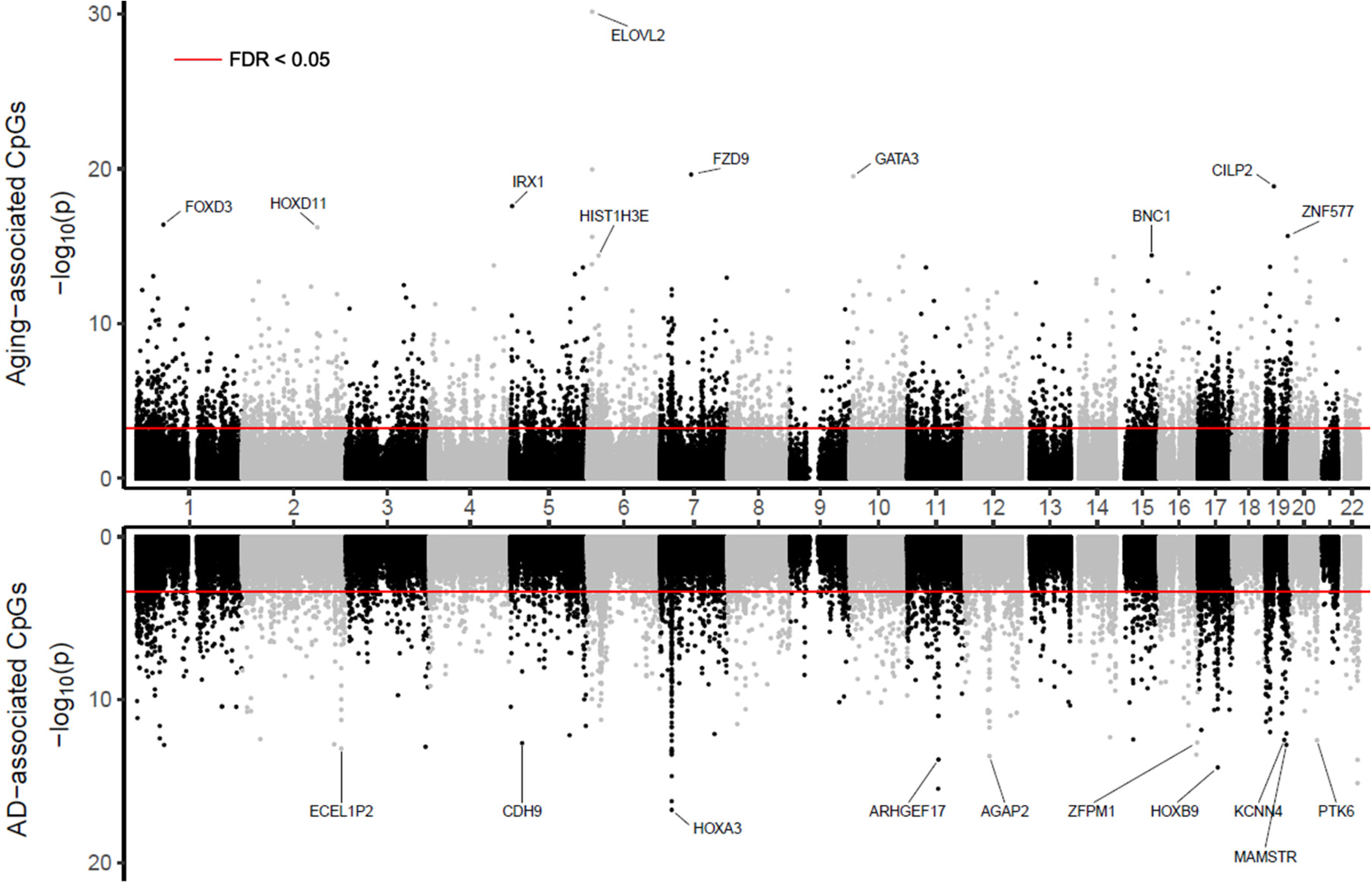
Miami plot of DNA methylation differences associated with aging and Alzheimer’s disease (AD) neuropathology in brain samples. The X-axis are chromosome numbers. The Y-axis shows -log_10_(*P-* value) of the CpG to age-at-death associations (above X-axis) or CpG to Braak stage associations (below X-axis). Red lines indicate significance threshold at 5% false discovery rate.

Among the top 10 most significant CpGs (Table 1), three CpGs, including the two most significant ones, are located in the promoter region of the *ELOVL2* gene, where DNA methylation in this region was shown to be one of the most robust markers of aging in various tissues, including both brain and blood ^48-50^. Interestingly, among the 3264 FDR-significant CpGs, 134 CpGs (4.10%), all of which are hypermethylated in aging, are located on genes belonging to the *HOX* family. This is significantly higher than the overall proportion of probes targeting *HOX* genes (1251 *HOX* gene probes among 740,555 tested probes, or 0.15%, *P-*value = 2.2×10^-16^). DNAm and gene expression changes in *HOX* genes have been reported during aging in various cell types, such as human hemopoietic cells, dermal cells, and muscle-derived cells^51-53^. The *HOX* genes encode transcription factors that determine cellular identity and ensure that stem cells can properly differentiate into various cell types needed to maintain tissue function and repair ^54,55^. These processes involve the continuous renewal, repair, and replacement of cells within a tissue to preserve proper tissue function.

In region-based analysis, after multiple comparisons correction, comb-p software ^34^ identified 191 significant DMRs associated with age at death at 5% Sidak adjusted *P-*value. Among them, 69 DMRs were also identified by the coMethDMR software ^35^ at 5% FDR (Supplementary Table 4). For these 69 DMRs, the average number of CpGs per DMR is 4.86 ± 6.23 CpGs. As with significant CpGs, the majority of the DMRs (86.96%, 60 DMRs) were also hypermethylated with increased age. Moreover, the most significant DMR is located in the *ELOVL2* gene, and five of the top 10 most significant DMRs are located in the *HOX* genes. Among other genes associated with the top 10 DMRs (Table 2), the *ISM1* gene encodes a secreted protein that is widely distributed across various body compartments and is involved in numerous biological processes in aging, including angiogenesis (formation of new blood vessels), metabolism, and immune responses ^56,57^. Our observed hypermethylation in the promoter region of the *ISM1* gene is consistent with a previous study that demonstrated *ISM1* gene expression decreases with age and nominated this gene as a potential biomarker for aging ^58^. The *KLF14* gene encodes a zinc finger-containing transcription factor. A recent study demonstrated that *KLF14* expression decreases with aging in various tissues, including blood, liver, and peripheral blood lymphocytes, and this decrease is associated with accelerated cellular senescence and aging-related pathologies ^59^. These findings are consistent with our observed hypermethylation in the promoter region of the *KLF14* gene associated with increased age.

**Table 2.**
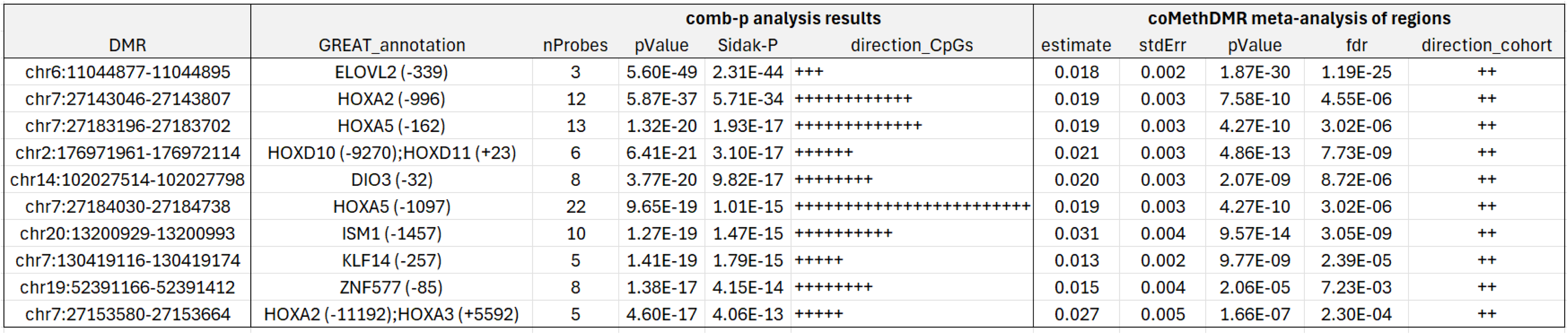
Top 10 most significant differentially methylated regions (DMRs) associated with age at death identified by both comb-p and coMethDMR software in meta-analysis. For each DMR, annotations include location of the DMR (DMR) and nearby genes based on GREAT. Comb-p results include the number of probes (nProbes), multiple comparison corrected P-value based on Sidak method (Sidak P), and direction of each CpG within the DMR (direction_CpGs). The results based on coMethDMR include estimated effect size (estimate) and standard error (stdErr), pValue, false discovery rate (fdr) from inverse-variance weighted meta-analysis regression model, and direction of effect size in each cohort (direction_cohort). All *P-*values are two-sided. In GREAT annotation, the numbers in parentheses indicate distance from the TSS.

### Enrichment analysis highlighted the potential functional relevance of the aging-associated DNA methylation differences

We next tested the enrichment of the aging-associated methylation differences against various genomic features. Both significant hypermethylated individual CpGs and DMRs were enriched in *CpG island*, but depleted in the *open sea*, *shore*, and *gene body* (Supplementary Table 5-6, Figure 2 a-b,). There were slight differences in the enrichment of hypermethylated CpGs and DMRs. The significant hyper-methylated individual CpGs were over-represented in *1^st^ exon*, and *TSS1500*, but under-represented in *TSS200* and *shelf*. On the other hand, the hyper-methylated DMRs were enriched in *TSS200*, but under-represented in *intergenic* regions. In contrast, both significant hypo-methylated individual CpGs and DMRs were enriched in the *open sea* but depleted in *CpG islands*. The hypo-methylated CpGs were additionally enriched in *shelf*, and *gene body*, and under-represented in *TSS200*, while the hypo-methylated DMRs were under-represented in *TSS1500*.

**Figure 2.**
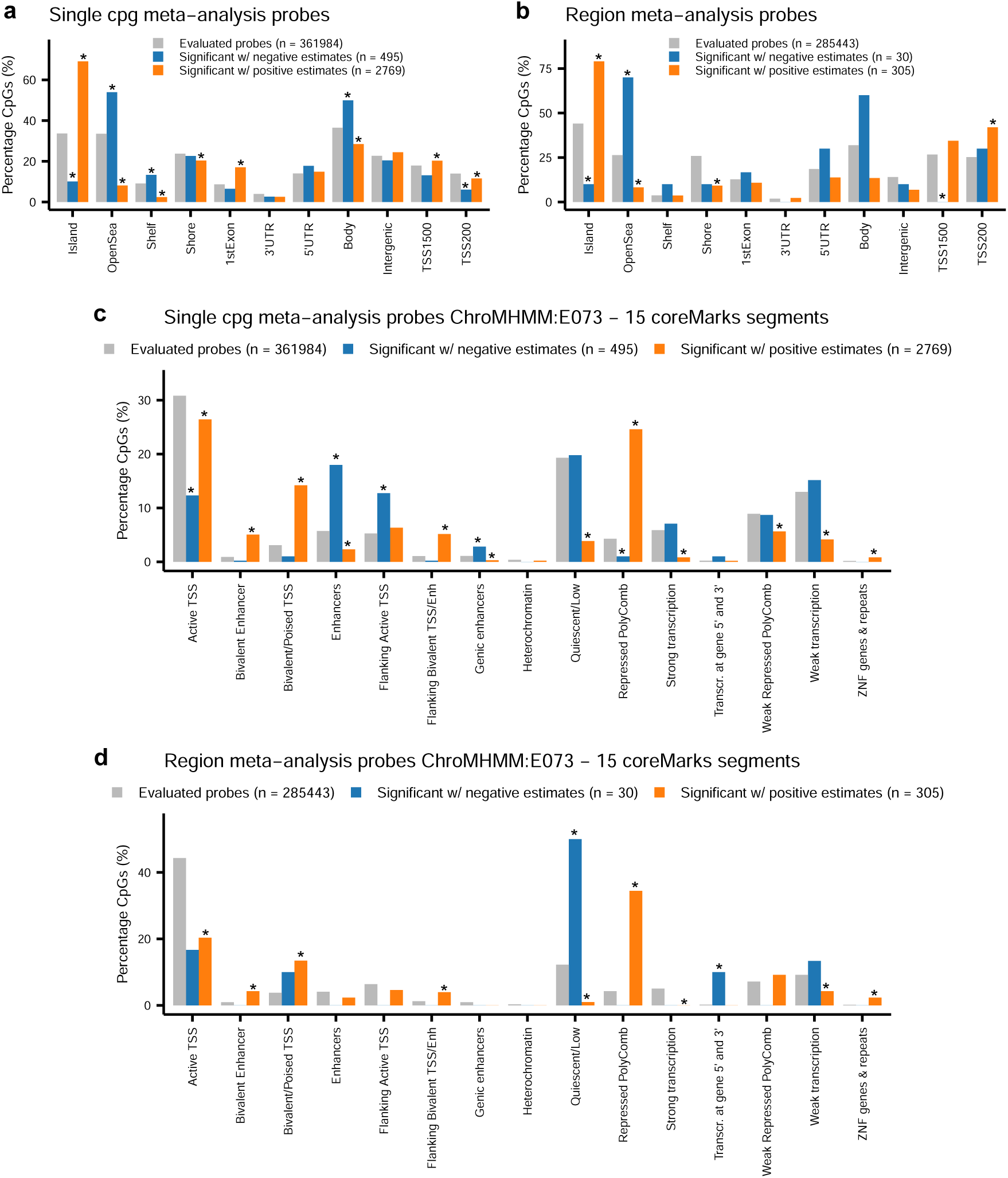
Enrichment of DNA methylation significantly associated with chronological age in meta-analysis of individual CpGs and DMRs. A two-sided Fisher’s test was used to determine over or under-representation of the significant CpGs in individual CpGs analysis and CpGs mapped within significant DMRs in various genomic features and chromatin states. Because a total of 26 genomic features (11 genomic locations and 15 chromatin states), we considered *P-*values less than 1.92 × 10^-3^ (i.e., 0.05/26) to be statistically significant.

In addition, we also compared our results with chromatin states generated by the Roadmap Epigenomics project^38,60^. Our enrichment analysis showed that significant hyper-methylated DMRs and CpGs were both enriched in regions with potential regulatory activity but are kept in a poised state (*bivalent enhancer*, *bivalent/poised TSS*, *flanking bivalent TSS/Enh*, *repressed polycomb*, and *ZNF genes & repeats*), but under-represented in regions with either active transcription (*active TSS*, *strong transcription*) or low level of transcription (*quiescent/low*, *weak transcription*) regions (Supplementary Table 7-8, Figure 2 c-d).

For hypomethylation in aging, there was less agreement for the enrichment of significant individual CpGs and DMRs. Specifically, the hypomethylated CpGs were enriched in *flanking active TSS* (Transcription Start Site)*, genic enhancers,* and *enhancers* which are distal regulatory elements that increase the transcription of target genes, but were depleted in *active TSS* and *repressed polycomb* regions. On the other hand, the hypo-methylated DMRs were enriched in *quiescent/low* and *transcription at gene 5’ and 3’* regions associated with transcription initiation and termination.

Similarly, enrichment tests for regulatory elements using the LOLA software^40^ also supported the potential functional relevance of these significant DNAm differences. In particular, the aging-associated DNAm were enriched in the binding sites of a number of transcription factors and chromatin proteins assayed by the ENCODE project^61^ (Supplementary Table 9). Notably, only age-associated hypermethylation, but not hypomethylation, was enriched in these regulators. Among the top hits were EZH2 and SUZ12, which are subunits of the polycomb repressive complex 2 (PRC2) protein complex. This finding is consistent with the observed enrichment of DNAm differences in PRC2 repressed regions and the CpG islands where PRC2 often binds (Fig. 2 c-d), as well as previous observations that DNAm often interacts with PRC2 binding^62-64^. We also found strong enrichment at binding sites of CTCF, a transcription factor essential for regulating gene expression and chromatin architecture by acting as an insulator. Intriguingly, aging-associated DNAm changes in blood have been consistently observed to be enriched at CTCF binding sites across multiple platforms for measuring DNA methylation ^12,65,66^. These DNAm at CpG sites within CTCF motifs can antagonize the binding of the CTCF protein, thereby affecting chromatin architecture and gene expression. It has been estimated as much as 40% of the variability in CTCF binding is attributed to DNAm ^67^. Other significant TFs and chromatin proteins also play important roles in aging by regulating various critical processes such as cellular metabolism (CtBP2, GR), development (CtBP2, Max, TCF12, TEAD4, NANOG), apoptosis (CtBP2, NRSF, Max), chromatin structure (CHD1, HDAC2, SETDB1), DNA repair (Rad21), and gene expression (HDAC2, SETDB1, TCF7L2, TEAD4, NANOG).

Finally, the gene ontology (GO) analysis of aging-associated DNA methylation differences highlighted the extensive influence of aging (Supplementary Table 10). Developmental processes, including embryonic and organ development, neural differentiation, and morphogenesis, are significantly enriched, indicating that aging impacts fundamental growth and differentiation pathways. The significant enrichment in cell fate specification and differentiation processes further suggests that aging is associated with the ability of cells to develop into specialized types, which is critical for tissue maintenance and repair. Additionally, the regulation of biological processes and communication, such as those involving blood circulation, points to age-related changes in physiological regulation and cell communication.

### Concordant DNA methylation differences associated with both aging and Alzheimer’s disease show a consistent direction of effect and common regulatory regions with enrichment

To further understand the methylation patterns in aging relative to Alzheimer’s disease, we compared the significant aging-associated CpGs and DMRs in this study (Supplementary Tables 3-4) with significant AD Braak stage-associated CpGs adjusting for age we previously identified in Zhang et al. (2020) ^19^ using the same DNA methylation processing pipeline and inflation correction procedures. We found that while the overwhelming majority (84.83%, 2769 out of 3264 CpGs) of aging-associated CpGs are hypermethylated, only slightly more than half (52.43%, 1451 out of 2767 CpGs) of the AD Braak stage-associated CpGs are hypermethylated. However, among the CpGs that are significant with both aging and AD neuropathology, almost all have concordant directions of change. Specifically, among the overlapping CpGs, almost all CpGs (249 out of 252 CpGs) that showed hypermethylation with increasing age also showed hypermethylation with advancing Braak stage (Figure 3a, Supplementary Table 11). Moreover, all CpGs (85 out of 85 CpGs) that showed hypomethylation with increasing age also showed hypomethylation with advancing Braak stage. Furthermore, among the 69 aging-associated DMRs, 15 are significantly associated with AD Braak stage, and all 15 DMRs have concordant direction of change in aging and AD Braak stage (Figure 3b, Supplementary Table 12). In Zhang et al. (2020), we also performed a matched samples analysis, in which each pathological AD case was paired with a control of the same sex and age at death within the same cohort, so that the DNAm differences from this analysis could be more confidently attributed to AD pathology rather than to confounding factors related to age or sex. The comparison of aging-associated CpGs and DMRs with AD Braak stage-associated DNAm in the matched analysis revealed a consistent pattern - all the DNAm differences significantly associated with both age (at death) and AD Braak stage showed concordant effect sizes in the same direction (Supplementary Figure 2, Supplementary Table 13 - 14).

**Figure 3.**
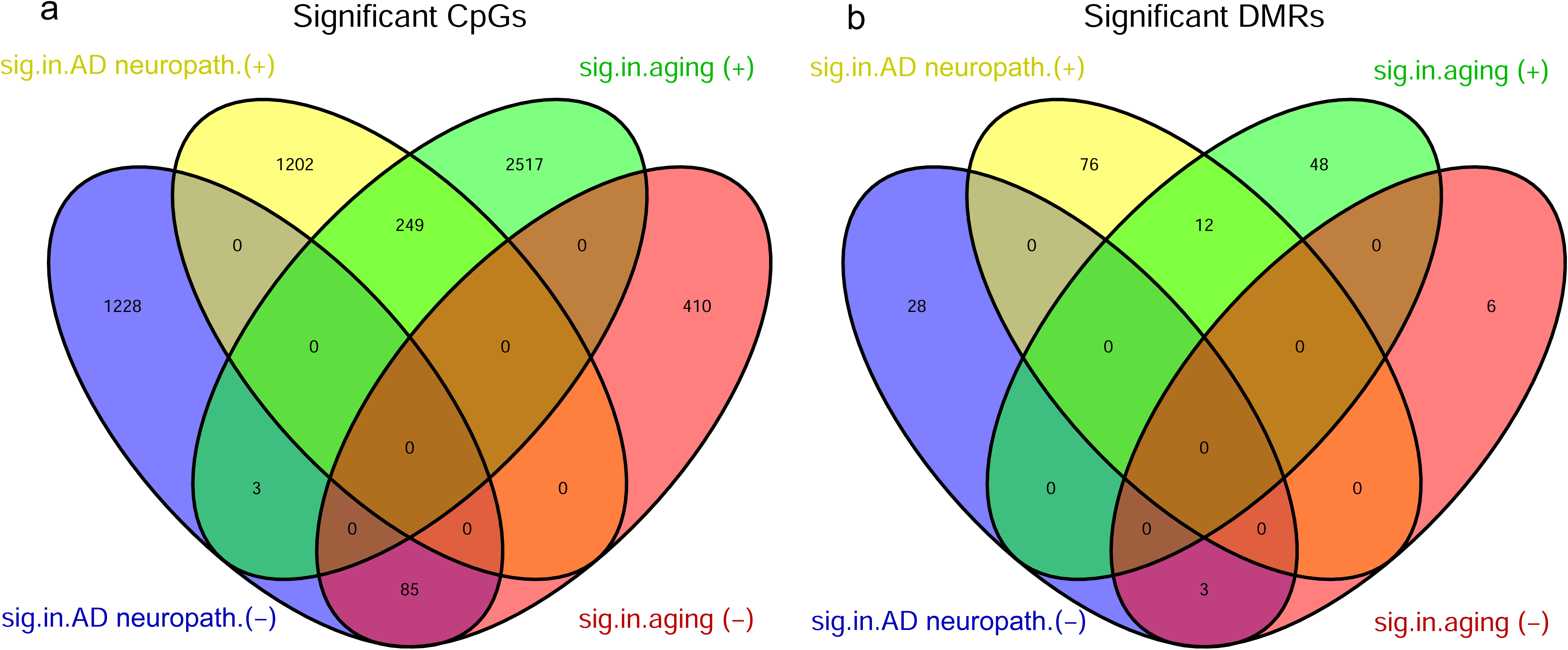
Almost all of the CpGs and DMRs significantly associated with both age and AD Braak stage showed concordant direction of change. In (a), among the overlapping CpGs, 249 out of 252 CpGs that showed hypermethylation with increasing age also showed hypermethylation with advancing Braak stage, and 85 out of 85 CpGs that showed hypomethylation with increasing age also showed hypomethylation with advancing Braak stage. Similarly, in (b), all 15 overlapping DMRs have concordant direction of change in aging and AD Braak stage. The significant AD Braak-associated DNAm were obtained from Supplementary Data 2 and 15 in Zhang et al. (2020) Nat. Comm. (PMID: 33257653). **Abbreviations** sig.in.aging (+): significantly hypermethylated in aging; sig.in.aging (-): significantly hypomethylated in aging; sig.in.AD neuropath. (+): significantly hypermethylated with increased AD Braak stage; sig.in.AD neuropath. (-): significantly hypomethylated with increased AD Braak stage.

In addition, we also compared enrichment analysis results of aging-associated DNAm with those of AD Braak-associated DNAm. Interestingly, we found a few discordant genomic features: while aging-associated CpGs are depleted in CpG island shores, gene bodies, and enhancers, AD Braak-associated CpGs are enriched in genomic regions with these features (Supplementary Table 5,7). However, the majority of the enrichment results are consistent and showed common regulatory regions. In both aging and AD neuropathology, the hypermethylated DNA methylation differences were enriched in CpG islands, but under-represented in open sea and shelf regions (Figure 2 a-b, Supplementary Table 5). Across the different chromatin states, in both aging and AD neuropathology, the hypermethylated DNAm differences were enriched in bivalent enhancers and repressed polycomb regions but depleted in active TSS and both quiescent/low and strong transcription regions (Figure 2 c-d, Supplementary Table 7). Taken together, these results highlighted the strong concordance in methylation changes (both hypermethylation and hypomethylation) between aging and AD neuropathology for DNAm that are significant in both analyses, underscoring common regulatory changes in both aging and AD.

### Co-localization analysis nominated genetic variants causal to both ADRD and aging-associated CpG methylation

To further understand the functional roles of the aging-associated CpGs and DMRs in AD, we next performed integrative analyses of aging-associated DNAm with GWAS summary statistics. First, to identify methylation quantitative trait loci (mQTLs) for the significant aging-associated DNAm differences, we searched for mQTLs for these DNAm loci in the brain xQTL database ^42^. Among the 3264 aging-associated CpGs (Supplementary Table 3) and the 335 CpGs located in significant DMRs (Supplementary Table 4), we found that 717 CpGs had 25661 mQTLs (Supplementary Table 15).

As aging and dementia may share common genetic factors, we next evaluated whether these mQTLs overlapped with genetic risk loci implicated in dementia, by comparing them with the genetic variants identified in a recent ADRD (Alzheimer’s and related dementia) meta-analysis ^43^. While no mQTLs overlapped with genome-wide significant loci for ADRD, we found 169 SNPs overlapped with genetic variants reaching a suggestive genome-wide significance threshold at *P* < 10^-5^ (Supplementary Table 16).

Given the observed overlap between the mQTLs and ADRD genetic risk loci, we sought to determine whether the association signals at these loci (variant to CpG methylation levels and variant to ADRD status) were due to a single shared causal variant or distinct causal variants in proximity. To this end, we performed a co-localization analysis using the method described by Giambartolomei et al. (2014)^68^. The results strongly suggested^69^ (i.e. PP3+PP4 > 0.90, PP4 > 0.8 and PP4/PP3 > 5) that 4 genomic regions included a single causal variant common to both phenotypes (i.e. ADRD status and CpG methylation levels). The CpGs associated with these causal variants are located in the *CHRNE*, *IDUA, KCNN4*, and *MLNR* genes (Supplementary Table 17).

### Brain-to-blood DNAm correlation analysis identified aging-associated brain DNAm with concordant cross-tissue changes

To identify aging-associated DNA methylation (DNAm) with the potential to serve as biomarkers, we next evaluated brain-to-blood DNA methylation correlations of the aging-associated DNAm. To this end, we utilized the London cohort dataset, which included 69 pairs of matched brain and blood samples^70^, and computed the Spearman correlations between brain and blood DNA methylation levels using an adjusted correlation analysis that accounted for confounding factors including estimated cell type proportions, batch effects, age, and sex in both brain and blood samples (see details in Methods).

Among the 3264 significant individual CpGs associated with aging and 335 CpGs located in aging DMRs, DNAm at 152 CpGs showed significant brain-to-blood correlations (*FDR* < 0.05) (Supplementary Table 18). Among the 152 CpGs with significant brain-to-blood correlations, all but one showed a significant positive association, corroborating previous analyses^70^ that observed a significant negative correlation between the brain and blood is relatively rare. The positive correlations ranged from 0.335 to 0.706. Notably, 22 (14.5%) of the 152 CpGs with significant brain-to-blood correlations are located on the *HOX* gene cluster (*HOXA2, HOXA4, HOXA5*) on chromosome 7. These regions with consistent cross-tissue epigenetic modifications may indicate important shared regulatory roles across tissues during aging.

### Validation of concordant brain DNAm differences in aging and AD using independent datasets

As mentioned above, we identified 249 CpGs hypermethylated in both aging and AD, and 85 CpGs hypomethylated in both conditions (Figure 3). To further validate these 334 (249 + 85) concordant brain DNAm differences, we examined their presence in previous AD studies using our recently developed MIAMI-AD database (https://miami-ad.org/)^71^. To this end, we made sure all referenced datasets were independent of ROSMAP and BDR. We found that among the 334 CpGs, 325 (97.3%) had nominal significant *P-*values in earlier studies, with consistent directions of change (Supplementary Table 19). Similarly, of the 15 DMRs with concordant aging and AD changes in Figure 3, all included one or more CpGs that reached nominal significance in previous analyses (Supplementary Table 20). Notably, many of these prior studies analyzed DNAm data from various brain regions (e.g., temporal cortex) or even whole blood, consistent with the observation that the majority of age-associated DNAm differences are not cell-type specific^66,72^. For example, PRC2-marked sites, where many of the aging-associated CpGs we identified are located, have been shown to undergo hypermethylation with aging, independent of tissue or cell type^73-76^.

### Concordant brain DNAm is associated with AD progression in out-of-sample validation

We next evaluated the feasibility of using concordant DNAm changes associated with both aging and AD as potential biomarkers. First, we overlapped the 334 CpGs with concordant DNAm changes with the 152 CpGs showing significant brain-to-blood correlations, which yielded 33 candidate CpGs. We then performed an out-of-sample validation using an external dataset from the ADNI study, which included 538 subjects with available DNAm data and follow-up visit information (Supplementary Table 21). For each subject, we analyzed the earliest available (baseline) DNAm sample measured in whole blood. These subjects were followed an average of 5.39 ± 2.94 years, with follow-up durations ranging from 0.44 to 11.71 years. By their last visit, 64 (30.0%) cognitively unimpaired (CU) subjects had progressed to mild cognitive impairment (MCI) or AD, and 131 (40.3%) MCI subjects had progressed to AD.

For each of the 33 candidate CpGs, we used Cox regression models to test whether baseline DNAm beta values were associated with disease progression (CU to MCI/AD or MCI to AD). The models were adjusted for age, sex, *APOE* ε4 allele status, years of education, and baseline MMSE score. At 5% FDR, we identified one CpG, cg10752406 located in the *AZU1* gene promoter, that was significantly associated with progression to the next disease stage (*P-*value = 0.0012, FDR = 0.0397) (Table 3). The Kaplan–Meier curve (Figure 4) shows that subjects in the highest and lowest tertiles of methylation for cg10752406 had significantly different survival probabilities over time. Although survival probability decreases in both groups over time, subjects with lower methylation consistently show a lower survival probability, indicating higher risk of AD progression. Moreover, hypomethylation at cg10752406 was significantly associated with both aging and increased Braak stage (Supplementary Table 22), and showed a significant brain-to-blood DNA methylation correlations (Figure 5), further supporting its potential as a peripheral AD biomarker.

**Figure 4.**
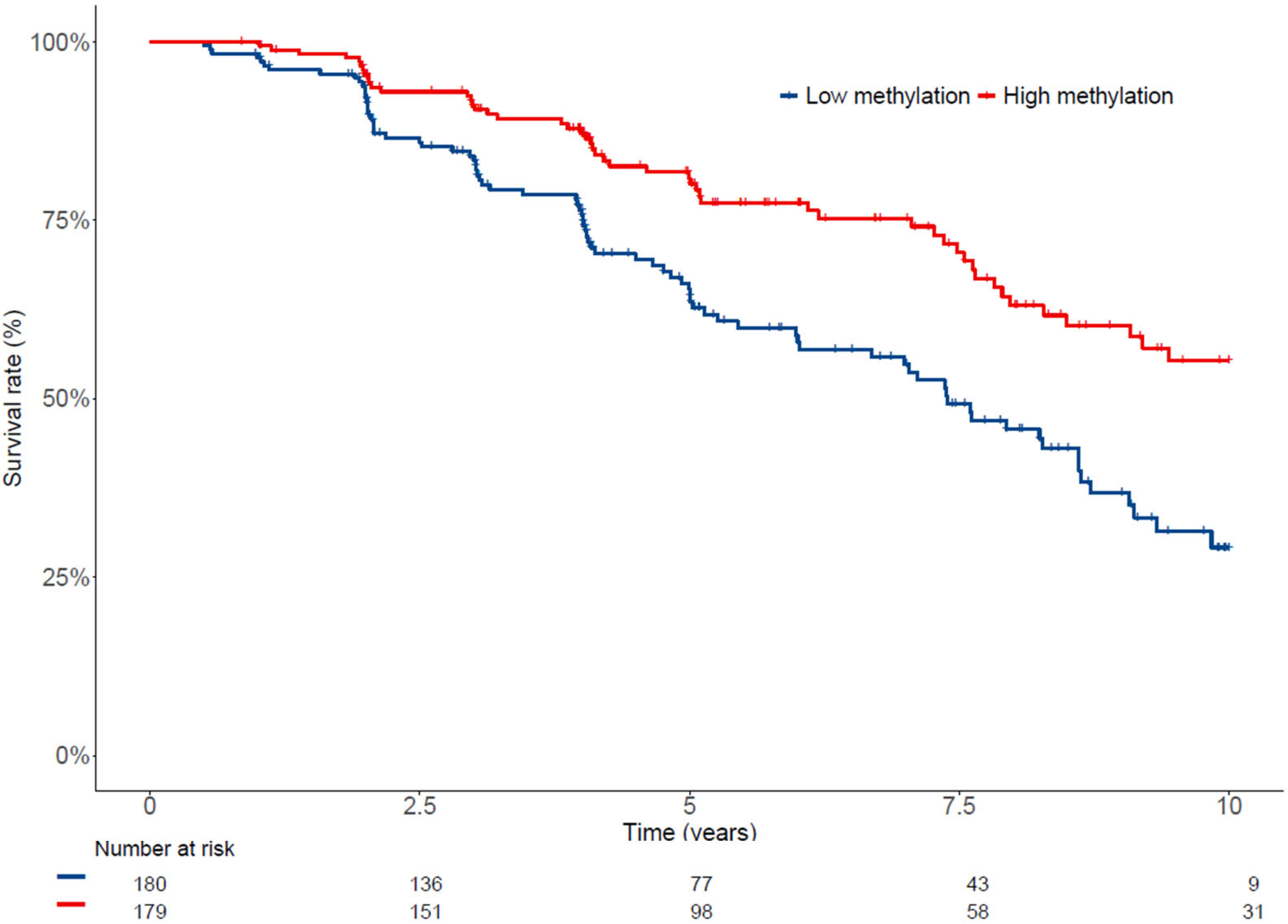
Kaplan-Meier curve for Alzheimer’s disease progression (CU to MCI/AD or MCI to AD) in the ADNI cohort, stratified by the highest and lowest tertiles of baseline DNA methylation at cg10752406. While survival probability declines in both groups, the low-methylation group consistently showed a lower survival probability, indicating a higher risk of AD progression. **Abbreviations** CU, cognitively unimpaired; MCI, mild cognitive impairment; AD, Alzheimer’s disease

**Figure 5.**
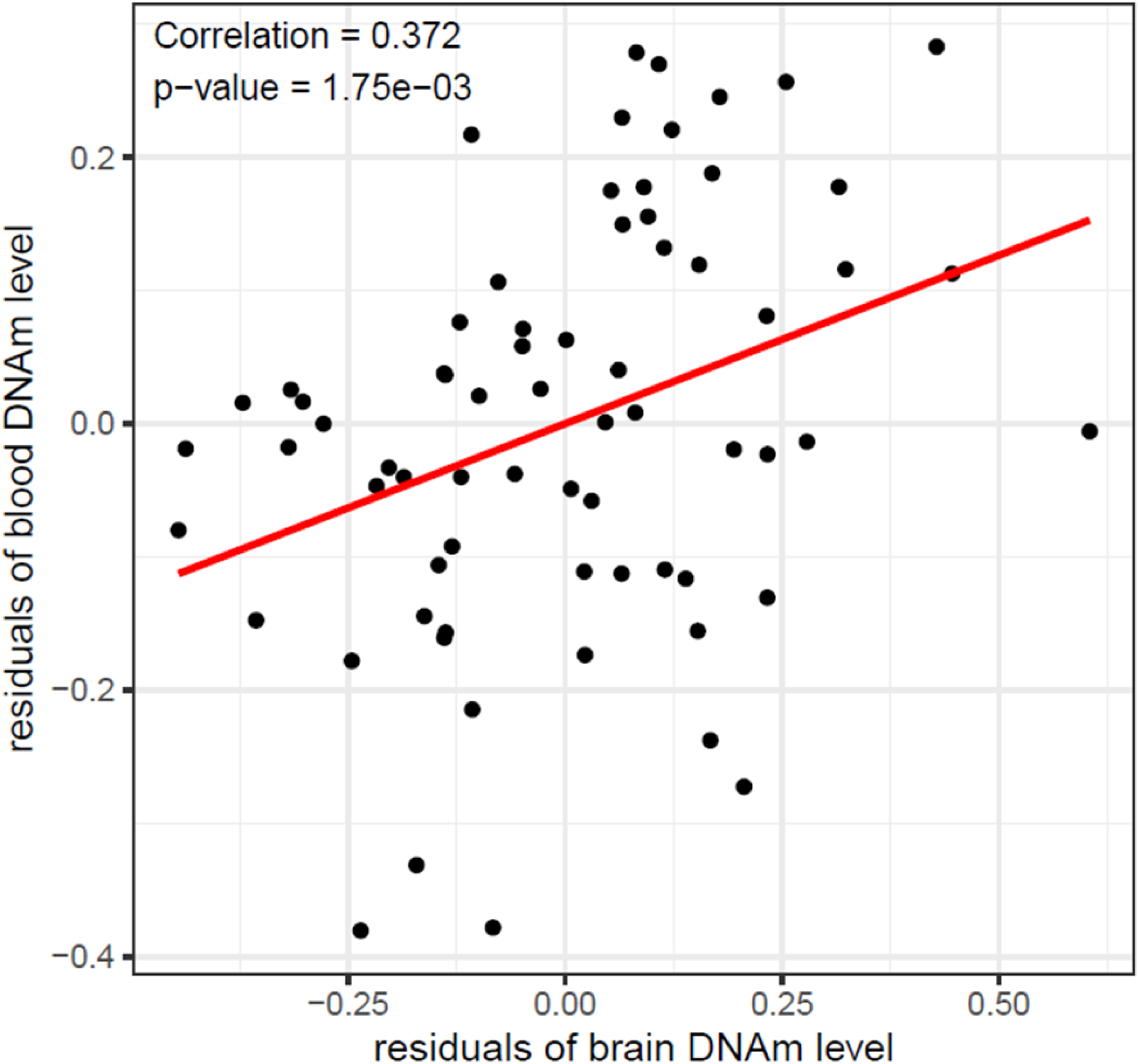
DNA methylation at cg10752406 showed significant brain-to-blood correlation in the London cohort samples (PMID: 26457534). Confounding effects were removed by first fitting linear regression models (methylation *M* value ∼ age + sex + batch + cell-type proportions) separately for brain and blood samples, and extracting the residuals. Spearman correlations were then computed using these residuals.

**Table 3.** Results from Cox regression model evaluating the association between DNA methylation at cg10752406 and disease progression (CN to MCI, MCI to AD) in 538 ADNI subjects, adjusted for age, sex, *APOE* ε4, MMSE, and education. Significant association was observed for cg10752406 (estimate =-5.853, *P-*value = 0.0012, FDR = 0.0397), indicating lower methylation at this CpG is associated with increased risk for AD progression.

| Model Term | Coefficient | HR (95% CI) | *P*-value |
| --- | --- | --- | --- |
| cg10752406 | -5.853 | 0.003 (0.0000831, 0.099) | 0.0012 |
| Age, years | 0.054 | 1.056 (1.034, 1.077) | $2.09 \times 10^{-7}$ |
| Male | 0.020 | 1.020 (0.752, 1.383) | 0.8992 |
| *APOE* ε4, allele count | 0.686 | 1.986 (1.596, 2.472) | $8.00 \times 10^{-10}$ |
| MMSE | -0.213 | 0.808 (0.742, 0.881) | $1.38 \times 10^{-6}$ |
| Education, years | -0.014 | 0.986 (0.932, 1.044) | 0.6369 |

**Abbreviations:** HR, hazard ratio; CI, confidence interval; MMSE, Mini-Mental State Examination

## DISCUSSION

We performed a comprehensive meta-analysis of two large cohorts of prefrontal cortex brain samples to prioritize consistent DNA methylation differences involved in aging. Consistent with previous brain sample studies ^46,47^, we found the majority of aging-associated DNAm differences were hypermethylated with increased age. Enrichment analysis showed that these hypermethylated aging-associated DNAm were significantly enriched in promoter regions and CpG islands, but under-represented in the open sea, shore, and gene body. In contrast, hypomethylated DNAm was enriched in open sea, shelf, and gene body regions, but depleted in CpG islands and promoter regions. Furthermore, comparison with chromatin state annotations generated by the Roadmap Epigenomics project ^38,60^ showed that hypermethylated aging-associated DNAm was enriched in genomic regions with potential regulatory activity but was maintained in poised states, such as bivalent enhancers and repressed polycomb regions. On the other hand, hypomethylated aging-associated DNAm were more likely to be found in distal regulatory regions, such as enhancers. Consistent with these findings, our integrative analysis with ENCODE data revealed hypermethylated DNAm, but not hypomethylated DNAm, were also enriched in binding sites of several transcription factors and chromatin proteins, including EZH2 and SUZ12, which are subunits of the PRC2 protein complex, as well as the insulator protein CTCF.

Consistent with a previous study ^46^, we found nearly all DNAm differences significantly associated with both age and AD Braak stage have concordant effect sizes. The geroscience view ^20^ posits that the biological processes involved in aging are critical in the onset and development of AD. It was proposed that aging and AD are intertwined - while molecular processes in aging can promote the disease, the disease can also influence the aging processes^20^. Relevant to DNA methylation, the Age-by-Disease Interaction hypothesis^77^ postulates that in many neuropsychiatric and neurodegenerative diseases, the trajectory of age-associated molecular phenotypes (e.g., gene expression) are modified and pushed in the disease-promoting direction and that DNAm is a potential mechanism contributing to the age-associated gene expression changes in AD^78^. Therefore, aging is a driving force rather than a confounding factor in the pathogenesis of AD.

Supporting this hypothesis, we found that many of the aging-associated DNAm differences identified in this study have also been previously implicated in AD. Among the 334 CpGs significantly associated with both aging and AD Braak stage, 97.3% had previously been linked to AD or AD neuropathology with consistent directional changes in prior studies. Importantly, these findings also pointed to new therapeutic targets. For example, we observed extensive hypermethylation at genes belonging to the HOX family. Previously, aberrant methylation on the *HOXA* gene cluster on chromosome 7 was shown to be associated with AD neuropathology in multiple AD EWAS datasets^22,79,80^. DNAm at the HOX genes has also been observed to interact with the protein complex PRC2, which regulates neuronal lineage specification and maintains neuronal functions. Importantly, PRC2 silences genes involved in neurodegeneration, and its deficiency leads to the de-repression of developmental regulators such as the HOX gene clusters, resulting in progressive and fatal neurodegeneration in mice^81^. Encouragingly, recent studies have demonstrated that rejuvenation can reverse epigenetic changes during aging. Increased physical activity, for example, has been observed to be associated with hypomethylation at *HOXA3* and *HOXB1* genes in human muscles ^53^.

Moreover, our out-of-sample validation using the ADNI dataset demonstrated that among the 334 CpGs with concordant brain DNAm differences in aging and AD, one CpG, cg10752406 located in the promoter of *AZU1*, was significantly associated with AD progression, even after controlling for age, sex, *APOE* ε4, years of education, and baseline MMSE score. This CpG also showed significant brain-to-blood DNAm correlations, further supporting its potential as a peripheral biomarker for AD. The *AZU1* gene, also called *CAP37,* encodes a neutrophil granule protein involved in inflammation and host defense^82^. In AD, the mRNA expression levels of *CAP37* is elevated, possibly due to increased amyloid beta and proinflammatory cytokines that activate microglial cells^83,84^. These findings underscore the significance of aging-related epigenetic modifications in AD progression and suggest that these DNAm sites may offer new opportunities for identifying high risk subjects for AD, thereby enabling earlier interventions.

This study has several limitations. First, we analyzed DNA methylation measured in bulk prefrontal cortex brain samples. To reduce confounding effects due to different cell types, we included estimated cell-type proportions as covariate variables in all our analyses. Although our results and previous studies have demonstrated that the majority of aging-associated DNAm is relatively robust across various tissues, future studies utilizing single-cell technology would provide more refined insight into the specific cell types affected by the aging-associated DNAm differences discovered in this study. Second, the Illumina arrays used by the studies analyzed in this meta-analysis cannot distinguish 5-methylcytosine (5mC) from 5-hydroxymethylcytosine (5hmC). Meta-analysis strategies similar to the one employed here could be applied to large datasets that discriminate between 5mC and further oxidized DNA modifications. Third, we focused our analyses on DNAm measured in the prefrontal cortex, a region significantly involved in aging and AD, partly due to the availability of large, high-quality DNAm datasets. It has been observed that there is a substantial correlation between DNAm in different brain regions ^85^. Moreover, previous studies have found the largest number of age-associated and AD neuropathology-associated brain DNAm changes in the frontal cortex compared to other brain regions (e.g., temporal cortex, entorhinal cortex, cerebellum) ^31,46,85^. Other brain regions, such as temporal cortex and entorhinal cortex, can be studied similarly as more data become available. Fourth, while Berger and colleagues identified aging-dysregulated histone modifications in AD ^86^, we did not observe similar DNA methylation patterns in our study. This discrepancy could be due to the limited coverage of the methylation arrays, which mainly focus on gene promoters and CpG islands. A previous study using next-generation sequencing technology found a number of hypomethylated DMRs associated with aging, with the majority lying outside of CpG islands and regions targeted by arrays^87^. Future studies with whole-genome bisulfite sequencing (WGBS) are needed to investigate the global landscape of aging DNAm in AD more comprehensively. Finally, we analyzed only DNAm samples from non-Hispanic white subjects. Future studies that investigate DNA methylation in large, multi-ethnic cohorts are needed.

In summary, we conducted a comprehensive meta-analysis to identify DNAm differences in the prefrontal cortex consistently associated with aging across two large brain sample cohorts. Our analyses revealed numerous robust DNAm differences associated with aging, highlighting key genes such as *ELOVL2*, *ISM1*, and *KLF14*, all of which have been implicated in various aging processes. We also observed significant overlaps between aging-associated DNAm changes and those involved in AD, supporting the hypothesis that aging and AD share common molecular underpinnings. Furthermore, our validation analyses using independent datasets confirmed both the reliability of the concordant brain DNAm differences in aging and AD and their potential to predict AD progression. Take together, these findings underscore the importance of aging as a critical factor in the pathogenesis of AD and suggest that age-related epigenetic modifications may offer a promising avenue for AD biomarkers.

## Data availability

All datasets analyzed in this study are publicly available. ROSMAP and BDR datasets can be accessed from AD Knowledge Portal (accession: syn3157275) and Gene Expression Omnibus database (accession: GSE197305). The London dataset used in integrative analysis can be accessed from Gene Expression Omnibus database (accession: GSE59685). The ADNI dataset can be accessed from http://adni.loni.usc.edu. The scripts for the analysis performed in this study can be accessed at https://github.com/TransBioInfoLab/AD-aging-brain-samples-analysis.

## Acknowledgment

Data used in the preparation of this article were obtained from the Alzheimer’s Disease Neuroimaging Initiative (ADNI) database (adni.loni.usc.edu). As such, the investigators within the ADNI contributed to the design and implementation of ADNI and/or provided data but did not participate in the analysis or writing of this report. A complete listing of ADNI investigators can be found at: http://adni.loni.usc.edu/wp-content/uploads/how_to_apply/ADNI_Acknowledgement_List.pdf

## Author’s contributions

L.W., J.Y., E.R.M., B.W.K designed computational analyses. D.L, L.G., M.A.S., W.Z., L.W. analyzed the data. L.W., J.Y., E.R.M, B.W.K., X.C. contributed to the interpretation of the results. L.W., D.L. wrote the paper, and all authors participated in the review and revision of the manuscript. L.W. conceived the original idea and supervised the project.

## ETHICS DECLARATIONS

### Ethics approval and consent to participate

The ADNI Study was approved by the Institutional Review Boards of all participating institutions. Written informed consent was obtained from all the participants or their authorized representatives. The study procedures were approved by the institutional review boards of all participating centers (https://adni.loni.usc.edu/wp-ontent/uploads/how_to_apply/ADNI_Acknowledgement_List.pdf), and written informed consent was obtained from all participants or their authorized representatives. The study was conducted in accordance with the Declaration of Helsinki and all study participants provided written informed consent for data collection. All work complied with ethical regulations for working with human participants. Ethics approval was obtained from the institutional review boards of each institution involved: Oregon Health and Science University; University of Southern California; University of California—San Diego; University of Michigan; Mayo Clinic, Rochester; Baylor College of Medicine; Columbia University Medical Center; Washington University, St. Louis; University of Alabama at Birmingham; Mount Sinai School of Medicine; Rush University Medical Center; Wien Center; Johns Hopkins University; New York University; Duke University Medical Center; University of Pennsylvania; University of Kentucky; University of Pittsburgh; University of Rochester Medical Center; University of California, Irvine; University of Texas Southwestern Medical School; Emory University; University of Kansas, Medical Center; University of California, Los Angeles; Mayo Clinic, Jacksonville; Indiana University; Yale University School of Medicine; McGill University, Montreal-Jewish General Hospital; Sunnybrook Health Sciences, Ontario; U.B.C. Clinic for AD & Related Disorders; Cognitive Neurology—St. Joseph’s, Ontario; Cleveland Clinic Lou Ruvo Center for Brain Health; Northwestern University; Premiere Research Inst (Palm Beach Neurology); Georgetown University Medical Center; Brigham and Women’s Hospital; Stanford University; Banner Sun Health Research Institute; Boston University; Howard University; Case Western Reserve University; University of California, Davis—Sacramento; Neurological Care of CNY; Parkwood Hospital; University of Wisconsin; University of California, Irvine—BIC; Banner Alzheimer’s Institute; Dent Neurologic Institute; Ohio State University; Albany Medical College; Hartford Hospital, Olin Neuropsychiatry Research Center; Dartmouth-Hitchcock Medical Center; Wake Forest University Health Sciences; Rhode Island Hospital; Butler Hospital; UC San Francisco; Medical University South Carolina; St. Joseph’s Health Care Nathan Kline Institute; University of Iowa College of Medicine; Cornell University and University of South Florida: USF Health Byrd Alzheimer’s Institute. The investigators within the ADNI contributed to the design and implementation of the ADNI and/or provided data but did not participate in analysis or writing of this report. A complete listing of ADNI investigators can be found online (http://adni.loni.usc.edu/wp-content/uploads/how_to_apply/ADNI_Acknowledgement_List.pdf).

### Consent for publication

Not Applicable

---

**Supplementary Figure 1.**
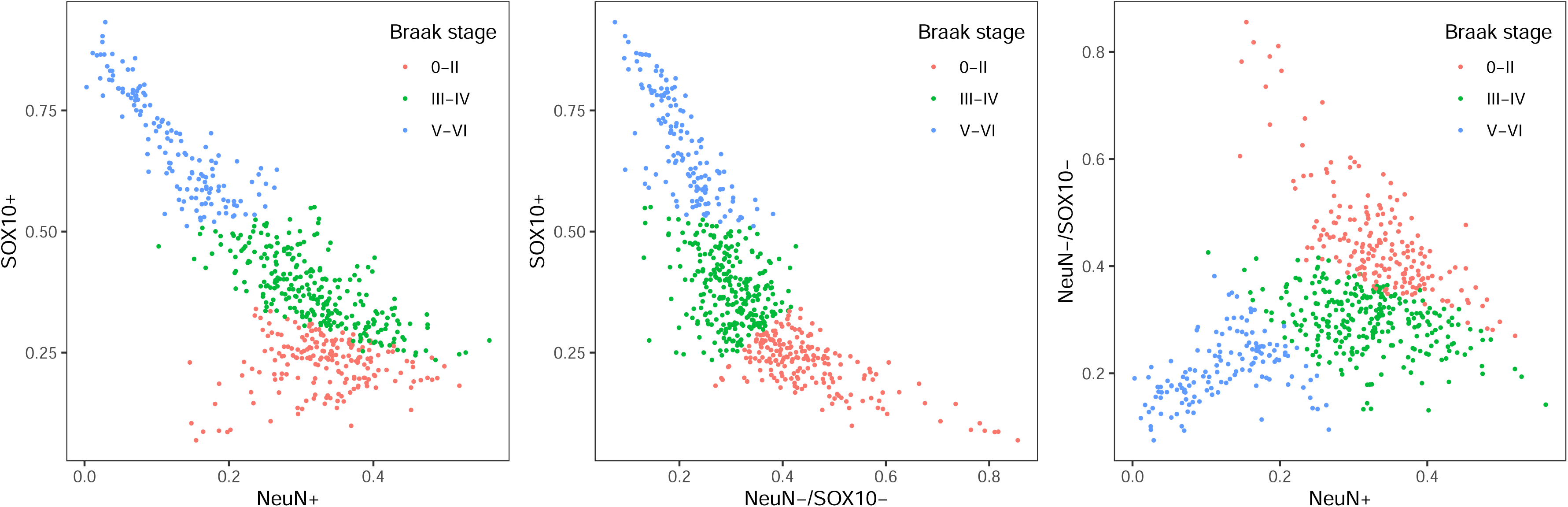
K-means (k = 3) clustering of bulk cortex brain samples in the BDR cohort using estimated relative cell type proportions of NeuN+, SOX10 +, and NeuN-/SOX10-. These results are consistent with those in Shireby et al. (2022) (PMID: 36153390), showing that an increase in tau pathology corresponds to a decrease in the proportion of NeuN+ (neuronal) cells, a reduction in NeuN-/SOX10- (microglial/astrocytic) proportions, and an increase in SOX10+ (oligodendrocytic) proportions in the brain cortex. The current study only used samples in the Braak stage 0-II group.

**Supplementary Figure 2.**
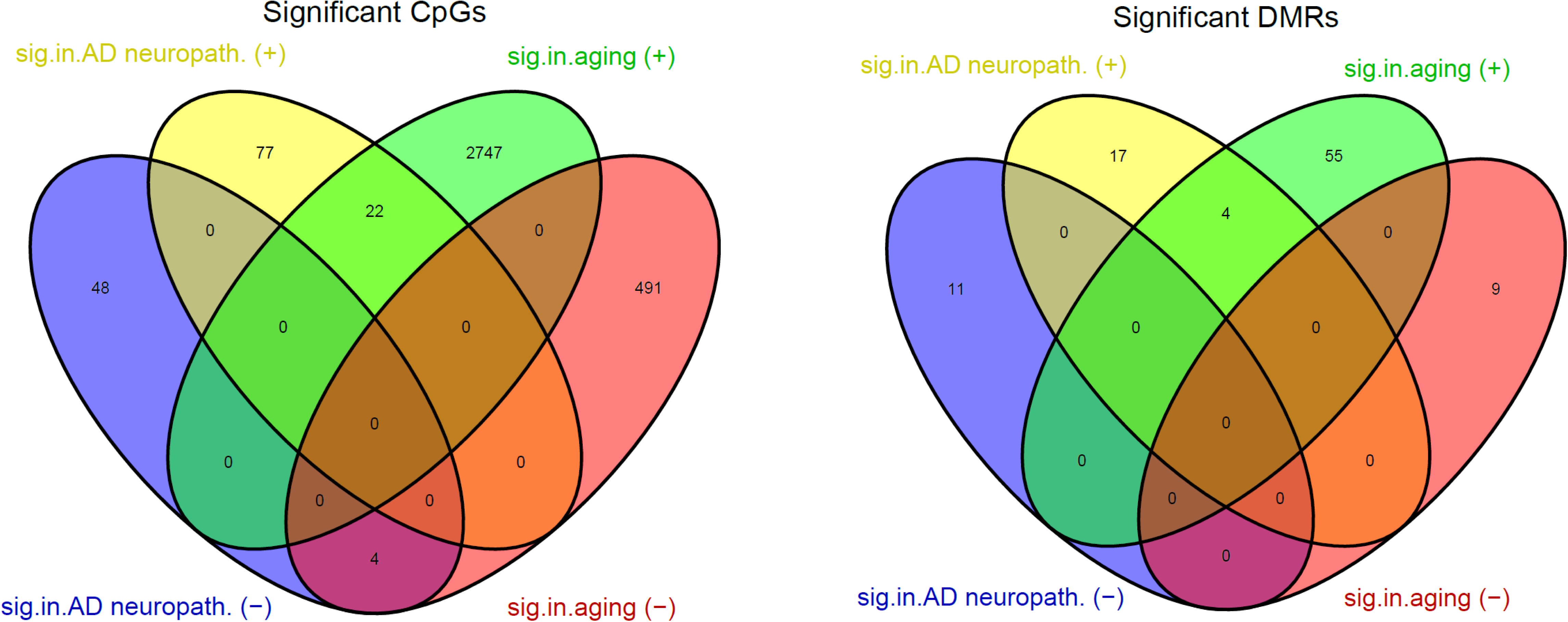
Comparison of aging-associated DNA methylation (DNAm) and AD Braak stage-associated DNAm (from matched samples analysis) shows that all of the CpGs and DMRs significantly associated with both age and AD Braak stage showed concordant direction of change. In (a), among the overlapping CpGs, 22 out of 22 CpGs that showed hypermethylation with increasing age also showed hypermethylation with advancing Braak stage, and 4 out of 4 CpGs that showed hypomethylation with increasing age also showed hypomethylation with advancing Braak stage. Similarly, in (b), all 4 overlapping DMRs have concordant direction of change in aging and AD Braak stage. The significant AD Braak-associated DNAm were obtained from Supplementary Data 6 and 7 in Zhang et al. (2020) Nat. Comm. (PMID: 33257653). **Abbreviations** sig.in.aging (+): significantly hypermethylated in aging; sig.in.aging (-): significantly hypomethylated in aging; sig.in.AD neuropath. (+): significantly hypermethylated with increased AD Braak stage; sig.in.AD neuropath. (-): significantly hypomethylated with increased AD Braak stage.

